# The Contrasting Role of Nasopharyngeal Angiotensin Converting Enzyme 2 (*ACE2*) Expression in SARS-CoV-2 Infection: A Cross-Sectional Study of People Tested for COVID-19 in British Columbia

**DOI:** 10.1101/2020.11.23.20237206

**Authors:** Aidan M. Nikiforuk, Kevin S. Kuchinski, David D.W. Twa, Christine D. Lukac, Hind Sbihi, C. Andrew Basham, Christian Steidl, Natalie A. Prystajecky, Agatha N. Jassem, Mel Krajden, David M Patrick, Inna Sekirov

## Abstract

**Background:** Angiotensin converting enzyme 2 (*ACE2*) serves as the host receptor for SARS-CoV-2, with a critical role in viral infection. We aim to understand population level variation of nasopharyngeal *ACE2* expression in people tested for COVID-19 and the relationship between *ACE2* expression and SARS-CoV-2 viral RNA load, while adjusting for expression of the complementary protease, Transmembrane serine protease 2 (*TMPRSS2)*, soluble *ACE2*, age, and biological sex.

**Methods:** A cross-sectional study of n=424 participants aged 1-104 years referred for COVID-19 testing was performed in British Columbia, Canada. Participants who tested negative or positive for COVID-19 were matched by age and biological sex. Viral and host gene expression was measured by quantitative reverse-transcriptase polymerase chain reaction. Bivariate analysis and multiple linear regression were performed to understand the role of nasopharyngeal *ACE2* expression in SARS-CoV-2 infection. The *ACE2* gene was targeted to measure expression of transmembrane and soluble transcripts.

**Findings:** Analysis shows no association between age and nasopharyngeal *ACE2* expression in those who tested negative for COVID-19 (P=0·092). Mean expression of transmembrane (P=1·2e-4), soluble *ACE2* (P<0·0001) and *TMPRSS2* (P<0·0001) differed between COVID-19-negative and -positive groups. In bivariate analysis of COVID-19-positive participants, expression of transmembrane *ACE2* positively correlated with SARS-CoV-2 RNA viral load (P<0·0001), expression of soluble *ACE2* negatively correlated (P<0·0001), and no correlation was found with *TMPRSS2 (P=*0·694). Multivariable analysis showed that the greatest viral RNA loads were observed in participants with high transmembrane *ACE2* expression (*B*=0·886, 95%CI:[0·596 to 1·18]), while expression of soluble *ACE2* may protect against high viral RNA load in the upper respiratory tract (*B*= −0·0990, 95%CI:[−0·176 to −0·0224]).

**Interpretation:** Nasopharyngeal *ACE2* expression plays a dual, contrasting role in SARS-CoV-2 infection of the upper respiratory tract. Transmembrane *ACE2* positively correlates, while soluble *ACE2* negatively correlates with viral RNA load after adjusting for age, biological sex and expression of TMPRSS2.

**Funding:** This project (COV-55) was funded by Genome British Columbia as part of their COVID-19 rapid response initiative.

## Research in Context

### Evidence before this study

We conducted a MEDLINE ^®^ search using the MeSH topic terms: “angiotensin converting enzyme 2 expression” and “SARS” and “age”, restricting the search to English-language reports published from January 1^st^, 2020. The search returned n=98 articles, eighty-eight of which reported primary research; these were further filtered by the MeSH qualifiers “epidemiology” and “virology” to provide n=43 articles. Search results were further restricted to the MeSH headings: age, virus replication, host pathogen interactions, nasal mucosa, RNA viral load and real-time polymerase chain reaction, which returned n=24 results. We read through the abstracts of these twenty-four papers and manually selected n=4 for full review. This review provided evidence that *ACE2* expression is greater in the upper respiratory tract than the lower respiratory tract, when measured by single-cell RNA sequencing, immunohistochemistry and high-sensitivity RNA in-situ mapping. In the upper airway, *ACE2* mRNA abundance closely correlates with protein concentration. A reverse genetics study demonstrated that a variable SARS-CoV-2 infection gradient occurs in the respiratory tract, with highest viral loads expected in the upper airway. In several patient cohorts, upper respiratory expression of *ACE2* was significantly increased in those who smoke, in this analysis multivariable adjustment of age suggested limited confounding.

### Added Value of this study

We measured *ACE2* expression in the context of COVID-19 testing to investigate the role of nasopharyngeal *ACE2* in SARS-CoV-2 infection. Our findings support previous work: the strong correlation we observe between nasopharyngeal *ACE2* expression and SARS-CoV-2 load also suggests an infection gradient across the human airway. Greater viral loads are expected in tissue with high transmembrane ACE2 expression. No observed relationship between age and nasopharyngeal *ACE2* in COVID-19-negative participants suggests that upper airway *ACE2* expression is independent of the RAAS pathway. We are the first to measure endogenous expression of soluble *ACE2* in the upper airway of people tested for COVID-19 and include this measure in multivariable analysis. Importantly, nasopharyngeal expression of soluble *ACE2* negatively correlates with viral RNA load, inferring a protective role at the population level.

### Implications of all the available evidence

Considering all available evidence, *ACE2* may play a dual, contrasting role in SARS-CoV-2 infection of the upper airway. Expression of transmembrane ACE2 positively correlates with SARS-CoV-2 RNA load, while expression of soluble ACE2 shows a negative association. Expression of nasopharyngeal ACE2 does not seem to correlate with age, as would be expected in the lower respiratory tract. Risk factors such as smoking may affect the baseline risk of high SARS-CoV-2 RNA loads in those infected. Factors associated with endogenous nasopharyngeal expression of soluble *ACE2* require further investigation.

## Introduction

In December 2019, clusters of viral pneumonia were reported in Wuhan, China. A novel highly pathogenic human coronavirus was isolated and named severe acute respiratory syndrome coronavirus 2 (SARS-CoV-2), the etiological agent of coronavirus disease (COVID-19).^1^ SARS-CoV-2 utilizes the same host receptor and protease for cell entry as the human coronaviruses SARS-CoV and HCoV-NL63. The receptor angiotensin converting enzyme 2 (*ACE2*) mediates cellular entry, while transmembrane serine protease 2 (TMPRSS2) serves as a complementary host factor.^2,3^ In human physiology, *ACE2* has a cardiovascular protective and anti-inflammatory role, as a constituent of the renin-angiotensin-aldosterone-system (RAAS).^4^ Interestingly, expression of *ACE2* in the nasopharyngeal tract exceeds that in alveolar tissue, explaining initiation of SARS-CoV-2 infection in the upper respiratory tract.^5^ Transcription of *ACE2* produces at least two dominant mRNA transcripts responsible for translation into soluble and membrane-bound protein isoforms.^6^ Although the transmembrane isoform has been shown to be crucial for viral entry into host cells, the role of soluble *ACE2* remains uncharacterized, though some evidence exists that it protects against SARS-CoV-2 infection.^7,8^ Transmembrane serine protease 2 (*TMPRSS2*) contributes to SARS-CoV-2 cell entry by cleaving the viral spike protein into a conformational form necessary for membrane fusion.^2^ Unlike *ACE2, TMPRSS2* expression occurs more stably across upper airway tissue and alternative enzymes such as cathepsin B/L may perform its role in viral infection interchangeably.^5^ To understand the importance of *ACE2* expression in SARS-CoV-2 infection, we performed a cross-sectional study of people tested for COVID-19 in British Columbia, Canada. The study aims to investigate the relationship between i) nasopharyngeal expression of *ACE2* and age in COVID-19-negative participants, ii) nasopharyngeal expression of host genes by COVID-19 test result and iii) nasopharyngeal expression of transmembrane *ACE2* and viral RNA load in those who tested COVID-19-positive adjusting for age, biological sex, expression of soluble *ACE2* and *TMPRSS2*. Characterizing expression of nasopharyngeal *ACE2* in a large group of participants tested for COVID-19 will increase our knowledge of host genes involved in viral infection and may allow for assessment of baseline risk.

## Methods

### Study Design and Participants

We performed cross-sectional sampling of people tested for COVID-19 at the British Columbia Center for Disease Control Public Health Laboratory (BCCDC-PHL) from 24/3/2020-9/5/2020. At the time of sampling, provincial health guidelines required a clinical indication for referral of a COVID-19 test. Inclusion criteria were applied to select study participants whose diagnostic specimens were: tested centrally at the BCCDC-PHL, collected by nasopharyngeal swab, the first test administered by provincial health number, suspended in Hologic Aptima™ media, negative for concurrent Influenza A, B or Respiratory syncytia virus infection, stored at - 80°C following RNA extraction and for whom host gene expression was successfully measured by quantitative reverse-transcriptase polymerase chain reaction (qRT-PCR). People meeting the inclusion criteria (n=444), were excluded (n=16) if samples had a SARS-CoV-2 E gene cycle threshold (Ct) value of >=38 by qRT-PCR. COVID-19-positive cases (n=212) were matched in a 1:1 ratio with those that tested negative for COVID-19 by age and biological sex (Figure S1).

Ethical approval for the study was obtained from the University of British Columbia human ethics board (H20-01110). Demographic variables of age and biological sex were drawn from public health laboratory data. Laboratory methods were performed in a College of American Pathologists accredited laboratory with externally validated qRT-PCR assays. ^9–12^

### Procedures

Nasopharyngeal samples collected in Hologic Aptima™ media were stored at 4°C before RNA extraction using the Viral RNA isolation kit on the MagMAX-96™ platform (ThermoFisher).^13^

Host and viral gene targets were assayed by qRT-PCR on the Applied Biosystems 7500 Real-Time PCR platform using TaqMan FastVirus 1-step polymerase (ThermoFisher). Total reaction volumes equaled 20*u*l, with 5*u*l of RNA template, 1*u*l of 20x primer/probe, 5*u*l Fast Virus and 9*u*l of nuclease free water per reaction. Cycling conditions were set to: 50°C for 5 minutes, 95°C for 20 seconds followed by 40 cycles of 95°C for 15 seconds and 60°C for 1 minute. A multiplex qRT-PCR reaction targeting SARS-CoV-2 envelope (E) and host ribonuclease P (RNaseP) was used to diagnosis acute viral infection by presence of viral RNA.^14^ Expression of RNaseP was used to ensure sample quality and control for sampling variation.^14^ Participants were diagnosed as COVID-19 positive with an E gene Ct value of <38. The E gene Ct values were transformed to genome equivalents per millilitre by making a 5-fold, 1:10 standard curve of SARS-CoV-2 synthetic RNA-MN908947·3 (Twist Bioscience) (Table S1). Commercially available primer probe sets were used to amplify the host gene targets TMPRSS2^12^ (Hs00237175_m1) (ThermoFisher) and *ACE2* ^10^ (Hs01085333_m1, HS01085340_m1) by multiplex qRT-PCR with a glyceraldehyde 3-phosphate dehydrogenase (*GAPDH*) control^11^ (Hs02758991_g1) (ThermoFisher). Relative gene expression was calculated between *TMPRSS2, ACE2* and *GAPDH* using the 2^-ΔΔCt^ method.^15^ Transmembrane *ACE2* was defined from the Hs01085333_m1 (ThermoFisher) gene target, which exon spans the transmembrane domain. Soluble *ACE2* was defined as absolute relative expression between the Hs01085333_m1 (ThermoFisher) and HS01085340_m1 (ThermoFisher) gene targets (Figure S3). The HS01085340_m1 target, does not include the transmembrane domain and is positioned between two motifs essential for SARS-CoV receptor binding (Figure S3).

### Statistical Analysis

De-identified data reporting participant COVID-19 test result, site of sample collection, type of collection media, accessioning laboratory, age and biological sex was accessed from public health laboratory records. Matching of COVID-19-positive people, who met the described inclusion, exclusion criteria, to COVID-19-negative participants by age and biological sex in a 1:1 ratio was performed by a nearest neighbor algorithm (n=424).^16^ Bivariate analysis was performed between: age, biological sex, viral RNA load, *TMPRSS2*, soluble or transmembrane *ACE2*, and COVID-19 test result. The balance of covariates between test groups was examined post matching.

The relationship between age and nasopharyngeal expression of transmembrane *ACE2* was examined in COVID-19-negative participants over the age of 18 by both linear regression and categorization of age into 9-year intervals. Differences in *ACE2* expression by age category were tested by analysis of variance.

Differences in mean expression of host genes by COVID-19 diagnosis was further examined by Levene’s test for equal variance and a two-tailed, t-test assuming non-equal variance.^17^ Correlation between host gene expression and viral RNA load was analyzed by simple linear regression. Multivariable analysis was performed by multiple linear regression, variable importance was assessed by the partial f-test.^18^ Collinearity was examined by the variable inflation factor with a cut-off of 10.^19^ The common cause criterion was applied to control for measured confounding, effect modification was incorporated when found statistically significant.^20^ All analysis was performed in RStudio version 1.2.5042 using the packages: car, ggsci, tidyverse, dataexplorer, ggpubr, lmtest, publish, forcats, matchit, tableone and effects. ^21^

### Role of the Funding Source and Data Stewards

The funder and data stewards played no role in the study design, analysis or interpretation of the results. As such, interpretation of the results does not reflect the views of the funding organization or data stewards. The study was performed as research at the University of British Columbia and British Columbia Centre for Disease Control.

## Results

The analytic dataset contains an age- and biological sex-matched sample of n=424 participants tested for COVID-19 in British Columbia from 24/3/2020-9/5/2020 (Figure S1). Participant characteristics are shown in Table 1. The mean age of n=212 COVID-19-positive participants was 61·6, 47·6% were biologically male. In n=212 COVID-19-negative participants, the mean age was 62·1, 48·6% were biologically male. Bivariate analysis between the examined covariates and diagnosis (COVID-19-negative or -positive) shows balance of age (P=0·83) and biological sex (P=0·92) between groups. Viral RNA (E gene Ct) was only detected in COVID-19 positive participants, with an average Ct value of 28·2. Relative expression of transmembrane *ACE2*, soluble *ACE2*, and *TMPRSS2* statistically differed by COVID-19 test result (Table 1).

**Table 1:** Structure and Characteristics of Analytic Data Stratified by COVID-19 Test Result (n=424).

| Variable Name | Level | Total (n) | COVID-19 Test Result |  | Chi X<sup>2</sup> | T-Test |
| --- | --- | --- | --- | --- | --- | --- |
|  |  |  | Negative | Positive |  |  |
|  | .. | 424 | 212 | 212 | .. | .. |
|  | .. |  | .. | .. | .. | .. |
| Age (mean [SD]) | .. | 424 | 62.2 [24.3] | 61.7 [23.5] | .. | 0.83 |
| Sex (n [%]) | .. | .. | .. | .. | 0.92 | .. |
|  | Male | 204 | 103 [48.6] | 101 [47.6] | .. | .. |
|  | Female | 220 | 109 [51.4] | 111 [52.4] | .. | .. |
| E Gene Ct (mean [SD]) | .. | 424 | .. | 28.2 [7.18] | .. | .. |
|  | .. | .. | .. | .. | .. | .. |
| Transmembrane ACE2 (mean [SD]) | .. | 424 | (0.00) [1.08] | (-0.609) [1.90] | .. | 1.2e-4 |
| Soluble ACE2 (mean [SD]) | .. | 424 | (0.00) [0.625] | (-0.898) [1.47] | .. | <0.0001 |
| TMPRSS2 Total (mean [SD]) | .. | 424 | (0.00) [0.797] | (-1.35) [1.60] | .. | <0.0001 |

Participants who tested negative or positive by COVID-19 were matched by age and biological sex, balance was checked between groups post matching by chi X<sup>2</sup> or t-test ( $\alpha=0.05$ ). Relative gene expression of *ACE2* and *TMPRSS2* was normalized to the negative group by the $2^{-\Delta\Delta C_t}$ method.<sup>15</sup>

### Relationship between ACE2 expression and age in COVID-19-negative participants

No relationship was found between nasopharyngeal *ACE2* expression and age in COVID-19 negative participants between 19 and 98 years of age with age as a continuous variable by linear regression (P=0·076). This finding was reproduced when age was categorized into nine-year intervals (P=0·092) (Figure 1).

**Figure 1:**
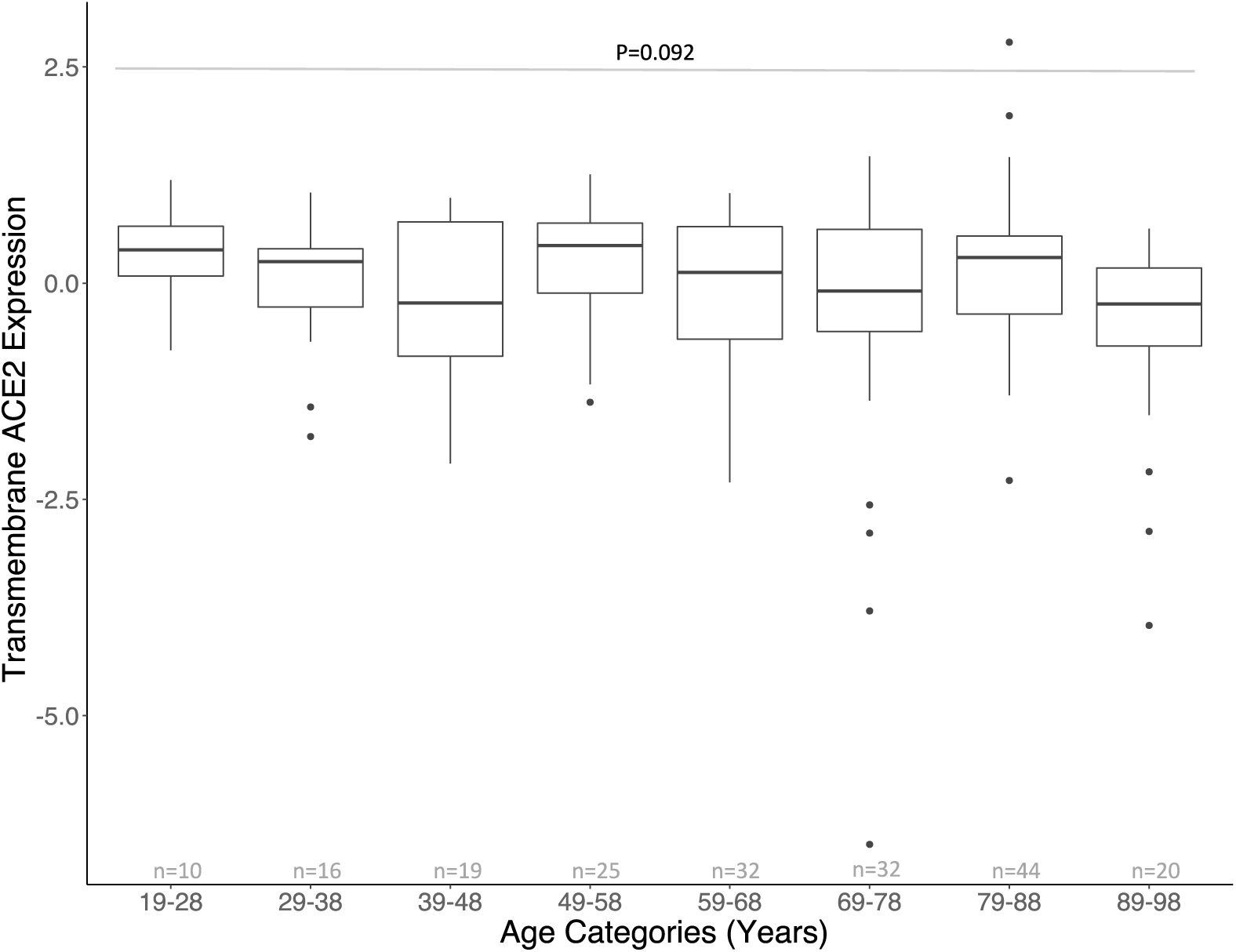
Relationship between age and nasopharyngeal transmembrane *ACE2* expression in COVID-19-negative participants. Boxplots of transmembrane *ACE2* expression by nine-year age categories in COVID-19-negative participants between the ages of 19 and 98 (n=198); boxes represent the Q1-Q3 interquartile range, whiskers represent 1.5x the Q1 or Q3 and horizontal lines the median transmembrane ACE2 expression by age category. Nine-year categories were selected to optimize the distribution of observations between groups. Participants who tested negative younger than 19 or older than 98 were excluded based on n<10 observations per age group. No difference was detected in mean transmembrane *ACE2* expression among age categories (ANOVA, P=0·092).

### Nasopharyngeal expression of ACE2 and TMPRSS2 by COVID-19 test result

Bivariate analysis showed a significant mean difference in transmembrane, soluble *ACE2* and *TMPRSS2* expression between COVID-19-negative and -positive participants (Figure 2). These differences were further examined to detect non-equal variance for all host gene targets by COVID-19 test result. Assuming non-equal variance, mean expression differed for transmembrane *ACE2* (P=1·2e-4), soluble *ACE2* (P<0·0001) and *TMPRSS2* (P<0·0001) between those testing COVID-19-negative or -positive (Figure 2). Expression of all three host genes was lower in COVID-19-positive participants.

**Figure 2:**
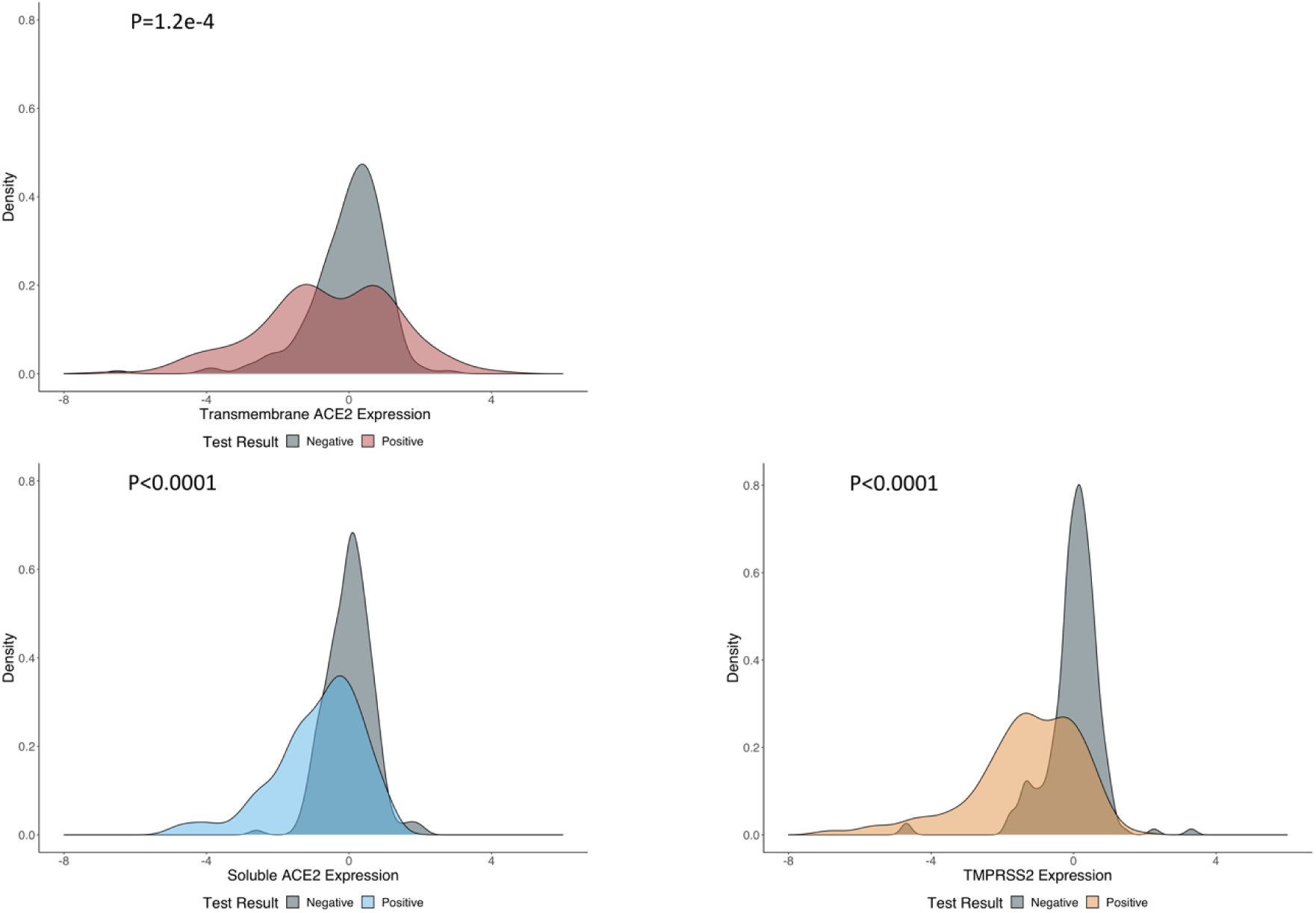
Relative nasopharyngeal gene expression of targeted host genes by COVID-19 test result. Gene expression is portrayed in kernel density plots stratified by COVID-19 test result. Probability densities of relative host gene expression are shown by positive (red, transmembrane *ACE2*; blue, soluble *ACE2*; yellow, *TMPRSS2*) and negative COVID-19 test results (grey). Levene’s test was used to detect non-equal variance in gene expression for all host targets between COVID-19-negative and -positive participants. A two-tailed, t-test was used to examine mean difference in host gene expression by COVID-19 test result assuming unequal variance: transmembrane *ACE2* (P=1·2e-4), soluble *ACE2* (P<0·0001) and *TMPRSS2* (P<0·0001).

### Association between host gene expression and viral RNA load in COVID-19-positive participants

Correlations between each host gene and viral RNA load were examined by simple linear regression. Nasopharyngeal expression of transmembrane *ACE2* positively correlated with viral RNA (P<0·0001). Expression of soluble *ACE2* negatively correlated with viral RNA (P<0·0001). No correlation was found between *TMPRSS2* expression and viral RNA (P=0·69) (Figure S2). Multiple linear regression estimated that a one-unit change in transmembrane *ACE2* expression increases viral RNA load by 0·886 Log_10_ GE/mL (95%CI: 0·596 to 1·18) adjusting for age, biological sex, expression of soluble *ACE2* and *TMPRSS2* (Table 2). No collinearity was detected between covariates. Biological sex could have been dropped from the model as evident from comparison between nested models (P=0·76), but was kept for validity. A partial f-test indicated effect modification between transmembrane and soluble *ACE2* expression (P=0·010) (Table 2). No effect modification was observed between transmembrane expression of *ACE2* and *TMPRSS2* (P=0·23). High expression of soluble *ACE2* decreases viral RNA load (*B*= −0·0990, 95%CI: [−0·176 to −0·0224]) (Table 2). The association between transmembrane *ACE2* expression and viral RNA in nasopharyngeal tissue differs by the concomitant level of soluble *ACE2* expressed (Figure 3). Effect modification was visualized by categorizing soluble *ACE2* by the relative mean expression +/− 2 standard deviations of all study participants (Low= −2·86, Mean= −0·444, High= 1·98, SD=2·43) (Figure 3).

**Table 2:**
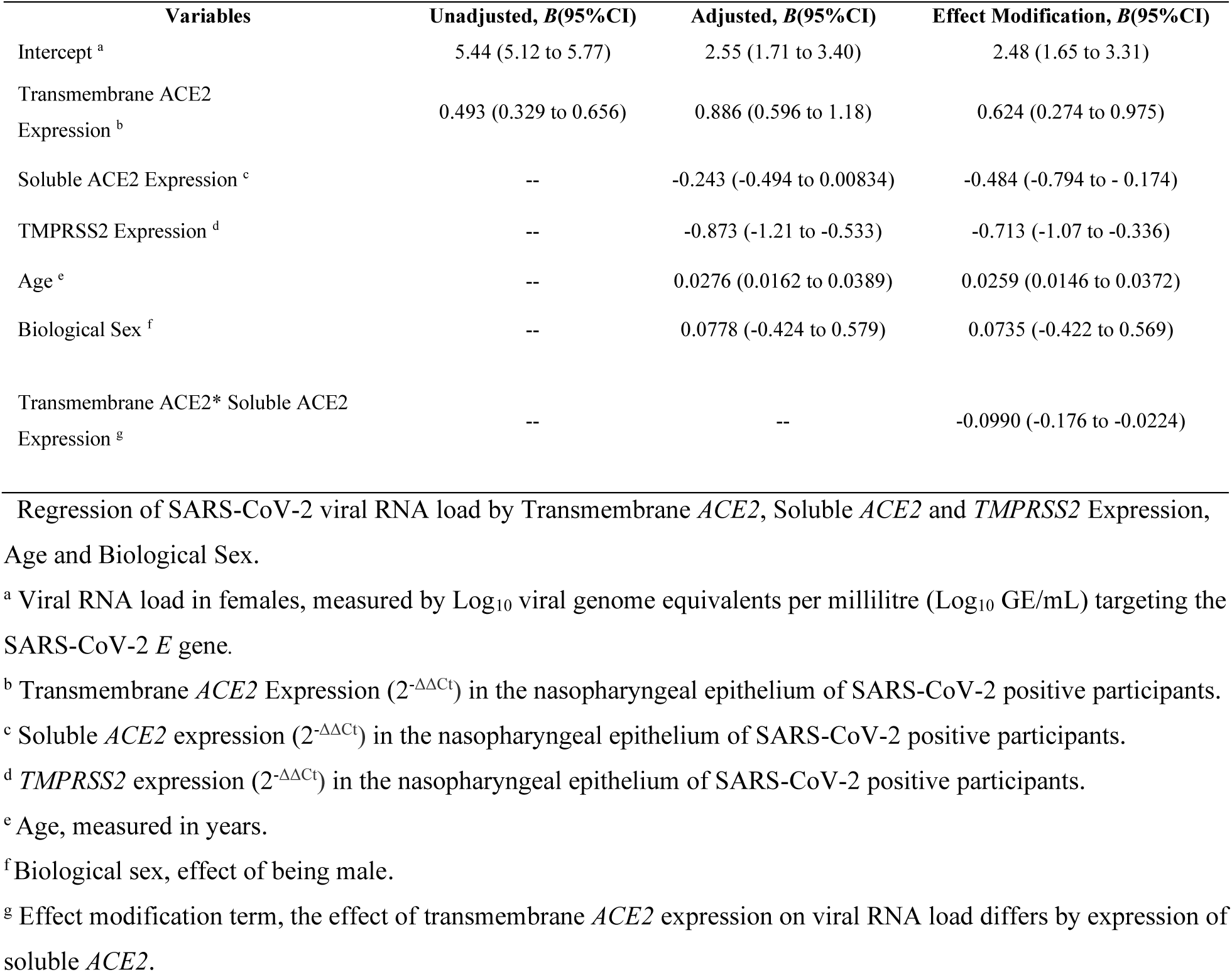
Unadjusted, Adjusted and Effect Modification Linear Regression Models of SARS-CoV-2 Viral RNA Load.

**Figure 3:**
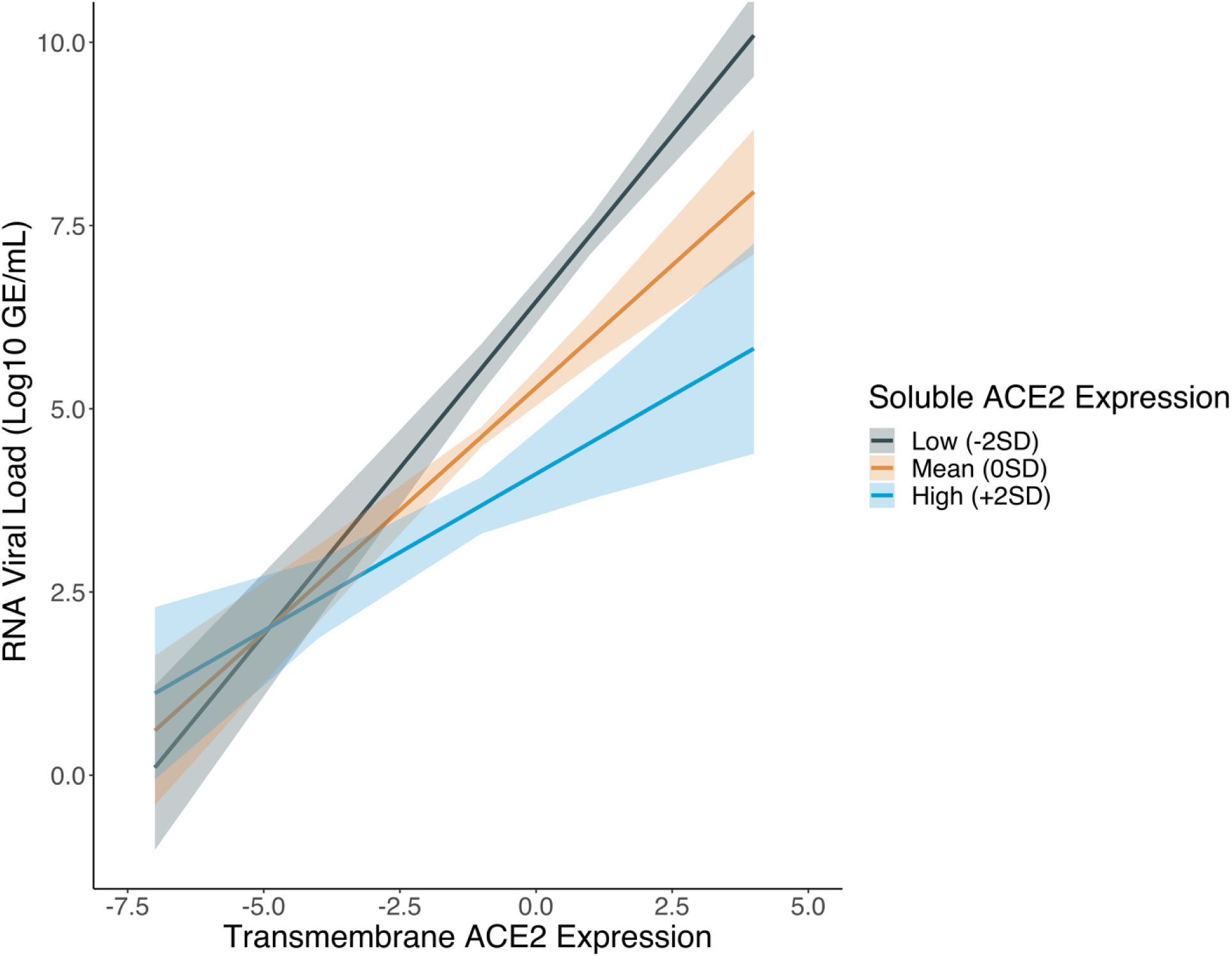
The association between transmembrane *ACE2* expression and SARS-CoV-2 RNA load in nasopharyngeal tissue differs by the amount of soluble *ACE2* expression. Soluble *ACE2* expression was categorized into low, mean and high levels to demonstrate the relationship. The low category (grey) represents soluble *ACE2* expression two standard deviations below the mean expression. The mean category (orange) codes for the mean soluble *ACE2* expression, zero standard deviations. The high category (blue) indicates soluble *ACE2* expression two standard deviations above mean expression. Shaded areas represent 95% confidence intervals, solid lines represent *B* coefficients from multiple linear regression.

## Discussion

In the described study, we measured nasopharyngeal *ACE2* and *TMPRRS2* relative gene expression by qRT-PCR in a cross-sectional sample of n=424 participants tested for COVID-19 in British Columbia. Analysis was performed to understand: the relationship between *ACE2* expression and age in COVID-19-negative participants, differences in host gene expression between COVID-19-negative and -positive participants and the role of *ACE2* in SARS-CoV-2 infection in those who tested positive for COVID-19.

Among participants tested negative for COVID-19, we found no relationship between nasopharyngeal *ACE2* expression and age, in a group of adults between 19-98 years of age. This result, agrees with findings from other efforts to characterize nasopharyngeal *ACE2* expression at the population level. Bunyavanich et al. reported a difference in *ACE2* expression levels between children younger than ten years old and young adults from eighteen to twenty-four years old. ^22^ No difference was evident between young adults, and adults twenty-five or more years in age. ^22^ Lack of a relationship between nasopharyngeal *ACE2* expression and progressive aging suggests that expression of *ACE2* in the upper airway does not correlate with expression of *ACE2* in the RAAS. The physiological importance of *ACE2* in the RAAS suggests that its expression would increase in response to age as older people have higher prevalence of cardiovascular disease or similar comorbidities (i.e. diabetes). ^23^ If nasopharyngeal expression of *ACE2* occurs independently of the RAAS pathway, future studies should ascertain what factors if any regulate expression of soluble *ACE2* in nasopharyngeal tissue and variation of *ACE2* expression in COVID-19-negative people over time. Some populations, such as young children, may have differential levels of nasopharyngeal *ACE2* expression which protect against SARS-CoV-2 viral infection.^22,24^

Participants who tested positive for COVID-19 were matched to those who tested negative by age and biological sex to estimate the direct relationship between host gene expression and SARS-CoV-2 test result. This analysis demonstrated that mean expression of transmembrane, soluble *ACE2* and *TMPRSS2* decreased in COVID-19-positive participants, while variation increased. Unfortunately, the limitation of reverse-causality in cross-sectional study design prevents us from understanding whether the presence of SARS-CoV-2 in positive samples affects host gene expression or host gene expression puts people at risk of viral infection. However, if we make the plausible assumption that at least a single round of viral replication must occur in order for qRT-PCR to generate a positive test result, it suggests that the observed variation in host gene expression of participants who test positive for COVID-19 results from viral replication.^25^ Numerous studies have demonstrated that coronavirus replication disrupts cellular transcription, as resources required by the cell to produce mRNA are instead sequestered by virus to replicate its genome. ^26^ Reduced transcription of *ACE2* by SARS-CoV, but not HCoV-NL63, in an *in vitro* model of Vero E6 cell infection was previously suggested as a pathological mechanism.^3^ Putative SARS-CoV-2 disruption of *ACE2* expression could partially explain the apparent association between hypertension, diabetes, and severe COVID-19.^27^ Answering these questions; however, requires a longitudinal study design to test the temporal effect of SARS-CoV-2 replication on the expression of *ACE2* and other implicated host genes.

The sample was then restricted to participants who tested positive for COVID-19 to investigate the association between nasopharyngeal *ACE2* expression and SARS-CoV-2 RNA load. Bivariate and multivariable analysis both suggest that *ACE2* plays a dual, contrasting role in SARS-CoV-2 infection. Transmembrane *ACE2* positively correlates with viral RNA load, while soluble *ACE2* may limit viral infection by reducing viral RNA load. Interestingly, effect modification was observed between transmembrane and soluble *ACE2* implicating that the proportion between these molecules may have more importance than absolute expression of either individually. For example, between two people with similar above-average nasopharyngeal transmembrane *ACE2* expression, we would expect a higher viral RNA load in the one with lower soluble *ACE2* expression. Though we are not equipped to characterize the mechanism by which soluble *ACE2* plays a protective role in SARS-CoV-2 infection, identification of this effect at the population level warrants further investigation of the underlying mechanism. Previous work has postulated that soluble *ACE2* restricts SARS-CoV/SARS-CoV-2 infection by acting as a decoy substrate.^28^ In a study of virus-*ACE2* dynamics in engineered human tissue, instead of binding to transmembrane *ACE2*, virus particles bound to soluble *ACE2* and were unable to infect susceptible cells.^28^ Moreover, recombinant soluble *ACE2* was recently administered to a COVID-19 patient requiring ventilation.^8^ In the case report, administration of soluble *ACE2* decreased viral RNA load in the patient’s plasma, tracheal aspirate and nasopharyngeal specimens. Additionally, soluble *ACE2* did not interfere with production of IgG and assumed its cardiovascular protective role in the RAAS.^8^

Soluble *ACE2* may also influence the usage of Neuropilin-1 as a co-receptor or alternative receptor for SARS-CoV-2. Function of Neuropilin-1 for cell entry and infectivity may depend upon high SARS-CoV-2 viral load; therefore, expression of soluble *ACE2* may limit SARS-CoV-2 tropism. ^29^

We acknowledge that the described study has several limitations. In our sample, the age distribution possesses a left skew which prevents us from examining the relationship between age and nasopharyngeal expression of *ACE2* in participants younger than 18 years of age. The difference in mean host gene expression observed between participants who tested negative or positive for COVID-19 cannot be resolved in a cross-sectional study design due to reverse causality. We attempted to control for immunological gene regulation by excluding people with endemic respiratory co-infections from Influenza A/B or Respiratory syncytia virus. We describe associations between transmembrane, soluble *ACE2* and SARS-CoV-2 RNA load which suggest the dual, contrasting role of *ACE2* in viral infection. However, we cannot provide evidence of the mechanism responsible for this association. Additionally, the measure of viral RNA load over-approximates infectious viral titre and viral RNA may be isolated in the absence of infectious virus.^30^

In conclusion, we have characterized nasopharyngeal *ACE2* expression in a sample of people tested for COVID-19. Analysis shows no relationship between age and nasopharyngeal *ACE2* expression in COVID-19-negative participants 19-98 years old. Expression of nasopharyngeal *ACE2* and *TMPRSS2* differs between COVID-19-positive and –negative groups. The role of nasopharyngeal *ACE2* in SARS-CoV-2 infection of the upper-airway may be differentiated by gene isoform expression; transmembrane *ACE2* positively correlates with viral RNA load, while soluble *ACE2* shows a negative association. Nasopharyngeal expression of *ACE2* possesses a dual, contrasting role in SARS-CoV-2 infection at the population level.

## Data Availability

The data that support the findings of this study are available from the data steward but restrictions apply to the availability of these data, which were used under license for the current study, and so are not publicly available. Data are however available from the authors upon reasonable request and with permission of the data steward.

## Contributors

AMN, DDWT, KSK, NAP, ANJ, MK, DMP, and IS conceived, designed, and obtained funding for the project. AMN wrote the manuscript, editing was provided by Karen Chu. AMN, DDWT and KSK performed the experiments. CDL and HS obtained and cleaned laboratory data. AMN, KSK, DDWT, CAB, CDL, and HS analyzed the data under the direction of CS, NAP, ANJ, MK, DMP and IS. All authors interpreted the data, edited the manuscript and provided their approval to publish.

## Acknowledgments

We would like to acknowledge the work of all our clinical colleagues at the British Columbia Centre for Disease Control and across the globe in responding to the COVID-19 pandemic. This work would not have been possible without funding from Genome British Columbia. We would like to thank Michael Donoghue for managing this multi-stakeholder project and Karen Chu for assisting as a medical copy editor.

## Supplementary Material

**Figure S1:**
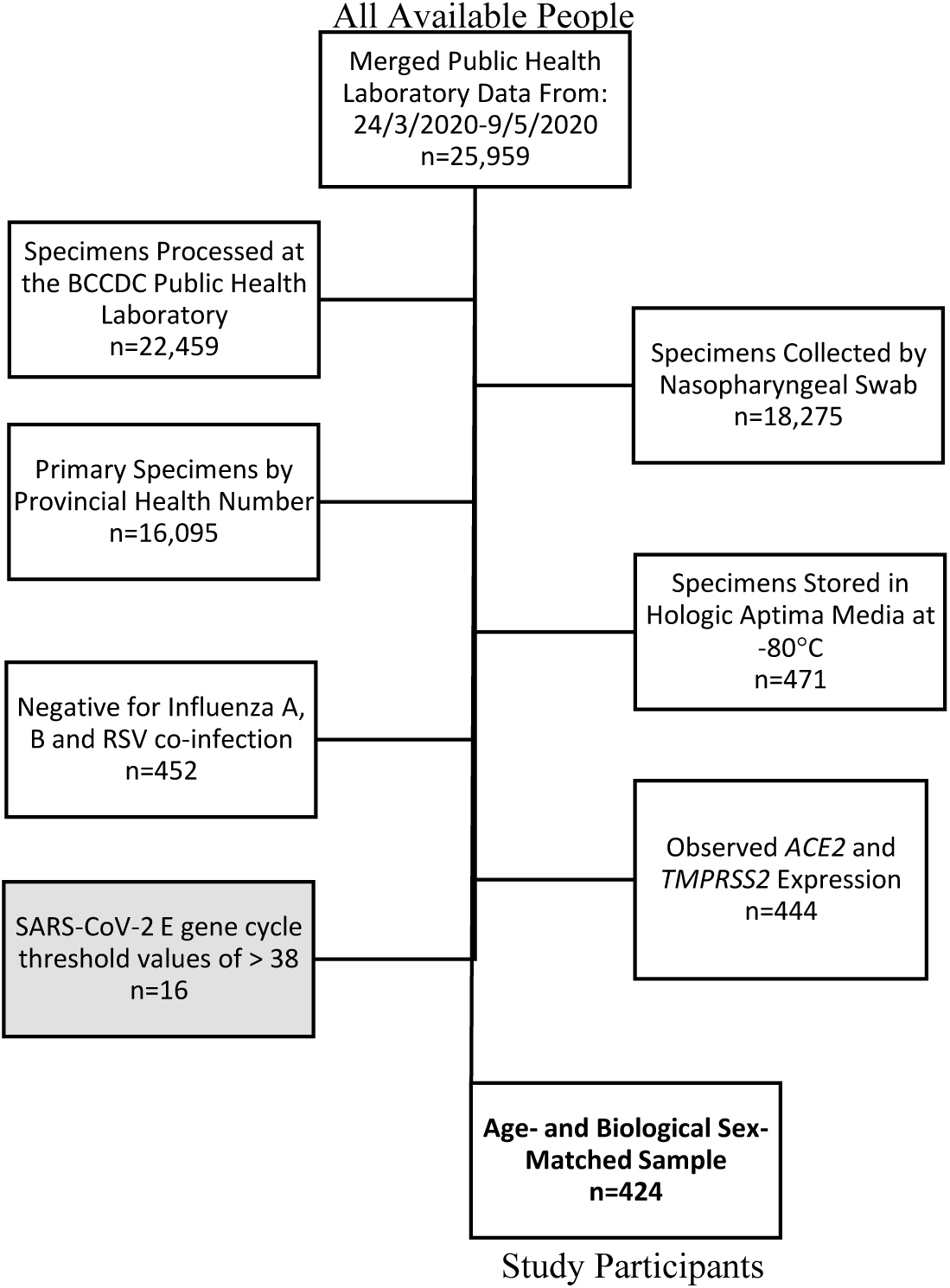
Inclusion, exclusion criteria applied to all people tested for COVID-19 in British Columbia from 24/3/2020-9/5/2020 to select study participants (analytic data). Specimens from people tested for COVID-19 with the following criteria were included (white box): tested at the BCCDC-PHL, specimen was collected by nasopharyngeal swab, test was the primary test by provincial health number, specimen was suspended in Hologic Aptima™ media and stored at −80°C following testing, negative for Influenza A, B or Respiratory syncytia virus infection and possessing host gene expression data. Those with a COVID-19-positive diagnosis, were further restricted by excluding (grey box) SARS-CoV-2 E gene Ct values of > 38 by qRT-PCR (n=212), and matched in a 1:1 ratio with COVID-19-negative people (n=212) by age and biological sex (bold text).

**Figure S2:**
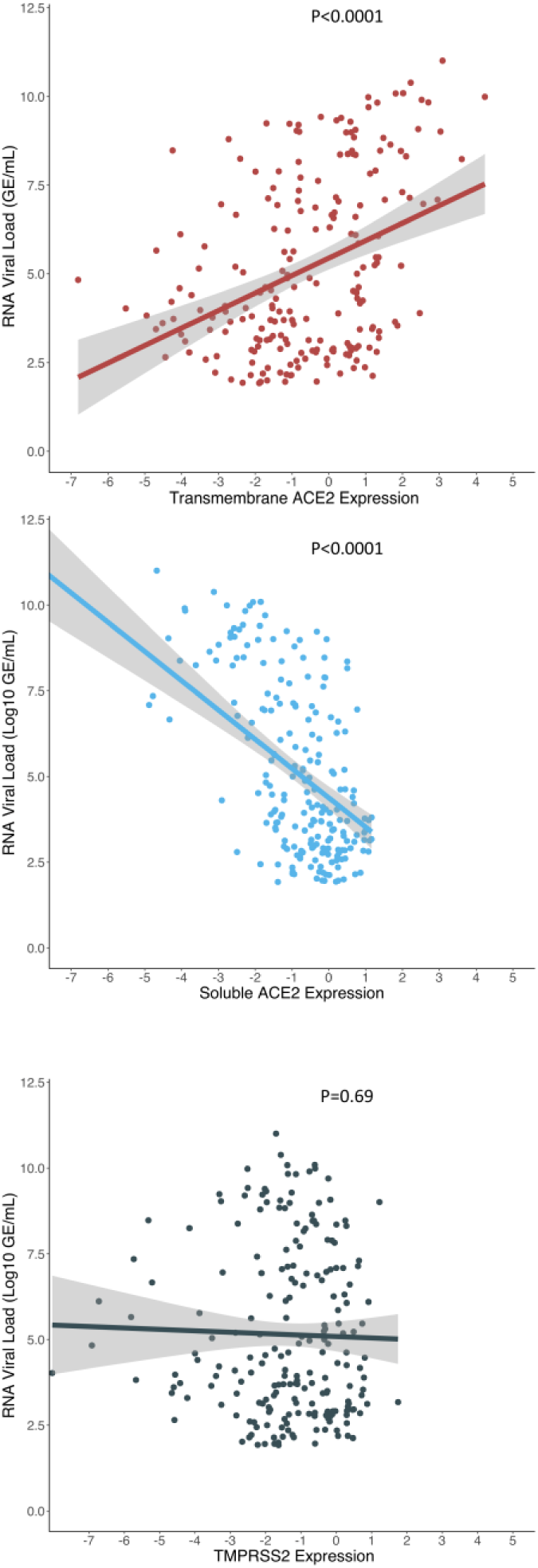
Correlation between host gene expression in COVID-19-positive participants and SARS-CoV-2 RNA load. The level of transmembrane *ACE2* (red), soluble *ACE2* (blue) and *TMPRSS2* (black) expression in positive cases of COVID-19 and its relationship with SARS-CoV-2 RNA is shown in Log_10_ genome equivalents per millilitre (GE/mL). Host gene expression was measured by quantitative real-time PCR and normalized to expression of the housekeeping gene GAPDH by the (2^-ΔΔCt^) method. Soluble *ACE2* was defined as absolute relative expression between the Hs01085333_m1 (ThermoFisher) and HS01085340_m1 (ThermoFisher) gene targets (Figure S3). Simple linear regression was used estimate correlations, the reported P values are for the *B*eta-coefficients of host gene expression.

**Table S1:** Serial Dilution of SARS-CoV-2 RNA for Generation of a Standard Curve by Quantitative Real-Time Polymerase Chain Reaction.

| **Log<sub>10</sub> Dilution** | **10<sup>0</sup>** | **10<sup>1</sup>** | **10<sup>2</sup>** | **10<sup>3</sup>** | **10<sup>4</sup>** |
| --- | --- | --- | --- | --- | --- |
| **~GE/mL<sup>‡</sup>** | **1** | **10** | **100** | **1000** | **10,000** |
| **R1** | 36.1 | 32.6 | 28.9 | 25.9 | 22.8 |
| **R2** | 35.8 | 31.9 | 28.8 | 25.8 | 22.8 |
| **R3** | 34.8 | 32.3 | 28.8 | 25.8 | 22.8 |
| **R4** | 35.6 | 31.7 | 28.8 | 25.7 | 22.8 |
| **R5** | 34.5 | 31.9 | 28.6 | 25.7 | 22.3 |
| **R6** | 34.0 | 32.1 | 28.7 | 25.5 | 22.7 |
| **R7** | 34.4 | 31.9 | 28.6 | 25.6 | 22.8 |
| **R8** | 34.1 | 31.4 | 28.8 | 25.6 | 22.8 |
| **R9** | 34.7 | 31.4 | 28.7 | 25.6 | 22.9 |

a 5-fold, 1:10 serial dilution of SARS-CoV-2 synthetic RNA was performed over nine independent replicates (n=9) to measure a standard curve by quantitative real-time PCR targeting the viral E gene.
<sup>‡</sup> a simple linear regression model was fit to n=9 replicates of the SARS-CoV-2 RNA standard curve: GE/mL= $b_0 + b_1x_1$ . Where, Genome Equivalents per millilitre (GE/mL) = 34.5 -1.27ln(x) and R<sup>2</sup>= 0.999.

**Figure S3:**
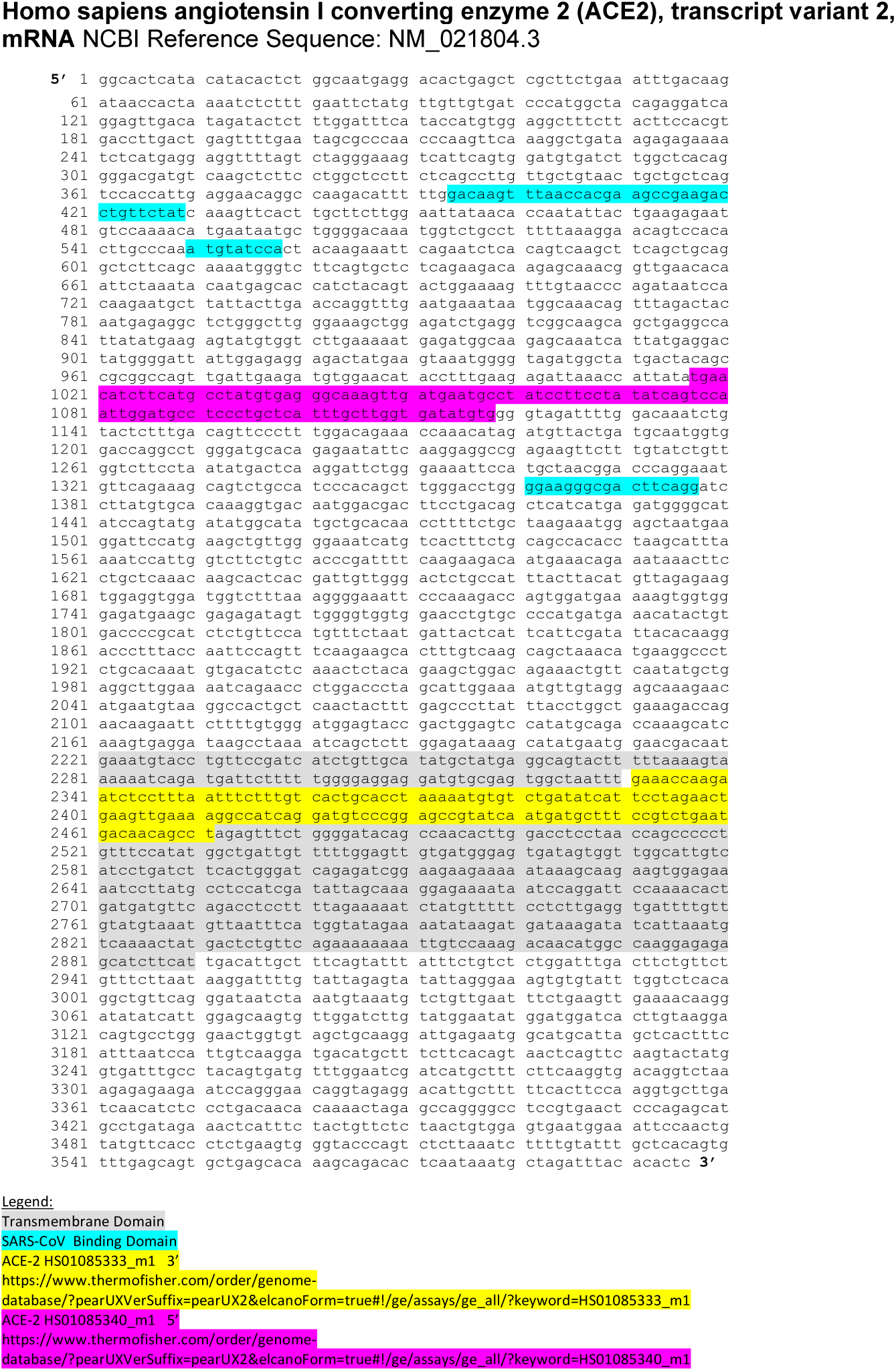
Nasopharyngeal expression of *ACE2* was determined by two quantitative real-time polymerase chain reaction targets, one target (yellow) exon spans the *ACE2* transmembrane domain. The other does not include the transmembrane domain (pink) and lays nested between two motifs known to interact with the SARS-CoV receptor binding domain (blue). The measure of soluble *ACE2* represents normalized expression of the pink to yellow target, or the proportion of non-transmembrane *ACE2* to transmembrane *ACE2*.

